# Disease and Social Media in Post-Natural Disaster Recovery Philippines

**DOI:** 10.1101/2021.03.22.21254137

**Authors:** Lauren E. Charles, Courtney D. Corley

## Abstract

**Introduction:** The Philippines is plagued with natural disasters and resulting precipitating factors for disease outbreaks. The developing country has a strong disease surveillance program during and post-disaster phases; however, latent disease contracted during these emergency situations emerges once the Filipinos return to their homes. Coined the social media capital of the world, the Philippines provides an opportunity to evaluate the potential of social media use in disease surveillance during the post-recovery period. By developing and defining a non-traditional method for enhancing detection of infectious diseases post-natural disaster recovery in the Philippines, this research aims to increase the resilience of affected developing countries through advanced passive disease surveillance with minimal cost and high impact.

**Methods:** We collected 50 million geo-tagged tweets, weekly case counts for six diseases, and all natural disasters from the Philippines between 2012 and 2013. We compared the predictive capability of various disease lexicon-based time series models (e.g., Twitter’s BreakoutDetection, Autoregressive Integrated Moving Average with Explanatory Variable [ARIMAX], Multilinear regression, and Logistic regression) and document embeddings (Gensim’s Doc2Vec).

**Results:** The analyses show that the use of only tweets to predict disease outbreaks in the Philippines has varying results depending on which technique is applied, the disease type, and location. Overall, the most consistent predictive results were from the ARIMAX model which showed the significance in tweet value for prediction and a role of disaster in specific instances.

**Discussion:** Overall, the use of disease/sick lexicon-filtered tweets as a predictor of disease in the Philippines appears promising. Due to the consistent and large increase use of Twitter within the country, it would be informative to repeat analysis on more recent years to confirm the top method for prediction. In addition, we suggest that a combination disease-specific model would produce the best results. The model would be one where the case counts of a disease are updated periodically along with the continuous monitoring of lexicon-based tweets plus or minus the time from disaster.

## Introduction

Located in Southeast Asia on the Pacific Ring of Fire, the Philippines consists of more than 7,000 islands, which are prone to meteorological (e.g., storms), hydrological (e.g., floods), and geophysical disasters (e.g., earthquakes and volcanoes). The latest intergovernmental panel on climate change (2014) reports with confidence that in Southeast Asia there will be a continued increase in 1) extreme weather conditions, involving strong variability in precipitation and increasing temperature, 2) compounded stress and adversity to sustainable development capabilities, 3) decrease in food production and security, 4) decrease in fresh water availability and security, and 5) rising sea level flooding affecting millions of people inhabiting the coastline [1]. Unfortunately, an increase in natural disasters and displacement of large proportions of the population combined with a decrease in food and water security will lead to a higher risk of communicable disease outbreaks after such disasters in developing countries in Southeast Asia [2].

To increase the Philippines’ population resilience, the developing country has enacted various emergency response programs, e.g., Surveillance for Post Extreme Emergencies and Disasters (SPEED), aimed at continuous monitoring of the affected populations during the emergency period [3, 4]. In these situations, evacuation centers are essential units used to house people for safety, provide medical attention, and issue disease surveillance. Hundreds of thousands of people can be relocated to these centers for two or more months depending on the severity, location, and type of disaster [5]. Despite coordinated efforts between local to global-level support, the centers take time to be set up properly with enough supplies to provide clean water sources and proper sanitation for the often overcrowded masses [6]. In response to Typhoon Haiyan, the majority of SPEED surveillance units were not operational within the recommended 48 hour window post-disaster [7]. Consequently, these periods of inadequate conditions are a perfect venue for communicable disease transmission in addition to the post-flooding environmental conditions creating an ideal habitat for disease vector population, e.g. mosquito, surges [8]. The result of both conditions lead to disease outbreaks even weeks to months after the original disaster occurred and, therefore, should play an important role in resilience-building strategies post-extreme weather events in the Philippines [9].

Currently, disease surveillance methods are effective only in controlling disease spread during the immediate aftermath of a disaster [10]. While social media posts, including Twitter data have been examined in the context of the immediate aftermath of a disaster [11], relatively little attention is invested to predict where disease outbreaks may occur after the emergency response phase, i.e., post-recovery. For developing countries, continuous monitoring and resources for health professionals, medical diagnostics, and disease surveillance is not available or practical, especially after a devastating natural disaster disruption. To help prevent disease outbreaks in high-risk areas, we need a method to increase the detection of disease post-natural disaster recovery, i.e., once the affected people are released from the evacuation centers. Previous research identifies social media as an informal source of near-real-time health data that may add valuable information to disease surveillance systems [11]. Social media provides broader access to health data across hard-to-reach populations [12]. This indirect health monitoring may improve public health professionals’ ability to detect disease outbreaks faster than traditional methods and to enhance outbreak response [13].

Coined the social media capital of the world, the Philippines provides a perfect opportunity to evaluate the potential of social media use in disease surveillance [14]. The objectives of this research are:

1. Determine the potential of publicly available Twitter data as an early warning of a likely communicable disease outbreak following a natural disaster in the Philippines
2. Given the data, compare various mathematical methods (e.g., statistical time series to machine learning neural networks) to identify the open-source algorithm that provides the best early warning capability to augment current disease surveillance capabilities.

By developing and defining a non-traditional method for enhancing detection of infectious diseases post-natural disaster recovery in the Philippines, this research aims to increase the long-term resilience of affected developing countries through advanced passive disease surveillance with minimal cost and high impact.

## Materials and Methods

This research was reviewed and approved by the Pacific Northwest National Laboratory (PNNL) Institutional Review Board.

### DATA

#### Twitter Data

We labeled geo-tagged tweets from the Philippines (2012-2013) based on the 17 regional boundaries and categorized them into the three island groups (Table 1). Filipino tweet language includes English, Tagalog, Taglish, and nearly 100 native dialects with locale-based cultural practices.

**Table 1:**
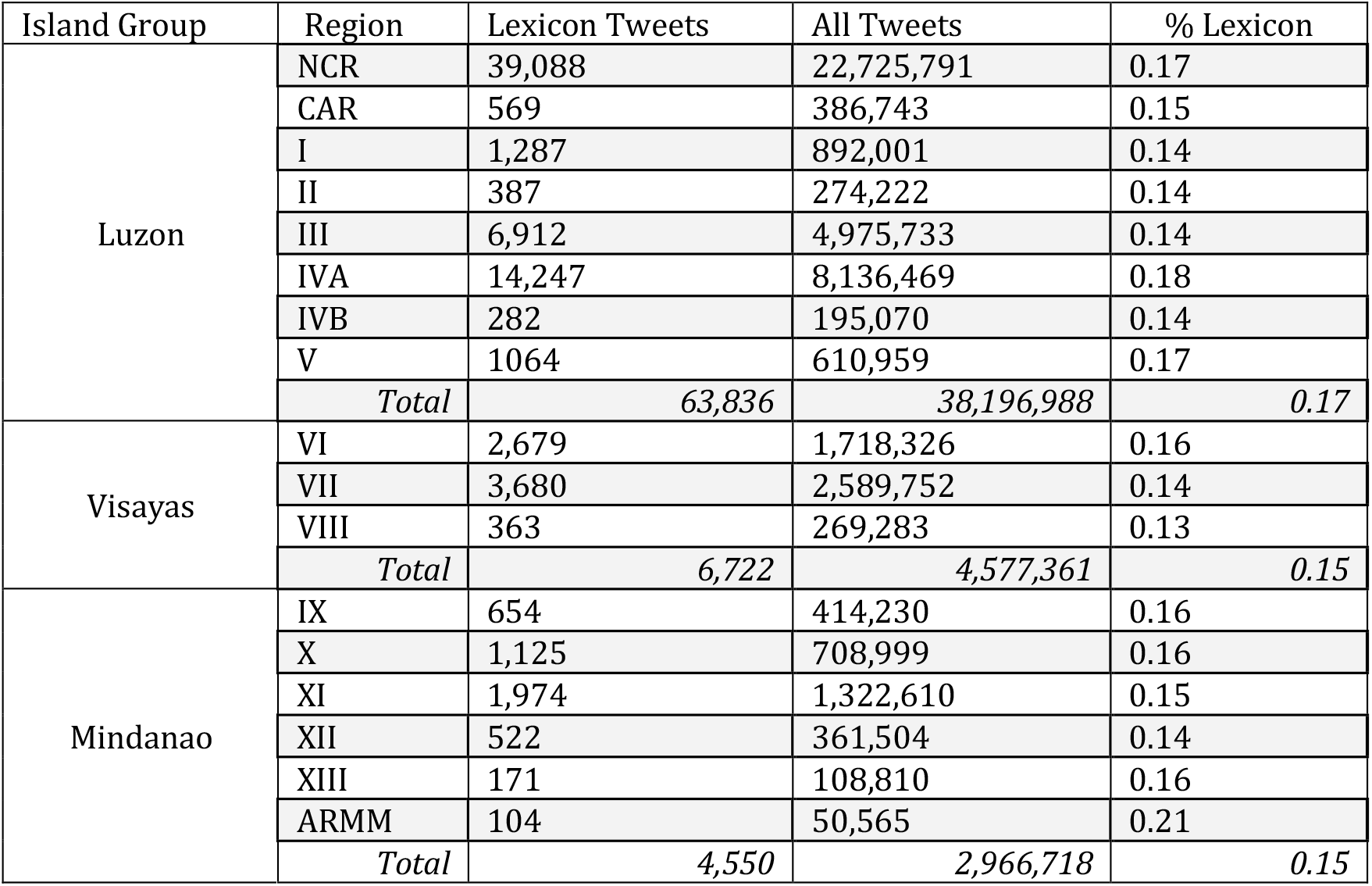
The total number of tweets collected from the Philippines, the number filtered by the disease lexicon, and the percent of tweets filtered by the lexicon shown by region and by Island group for all of 2012 and 2013.

#### Selected Disease Data

Disease case counts per week per region were obtained from the Philippines Department of Health for the following infectious diseases: cholera (CHOL), dengue (DEN), influenza-like illness (ILI), leptospirosis (LEPT), measles (MEA), and typhoid (TY).

#### Natural Disaster Data

The date, regional location, and type of natural disaster that occurred in the Philippines during 2012 and 2013 were collected from the websites of the Nationwide Operational Assessment of Hazards (NOAH; http://noah.dost.gov.ph/#/) and the Emergency Events Database (EM-DAT; www.emdat.be).

## ANALYSIS

### Disease Lexicon

A lexicon for sickness specific to the Philippines was devised based on the 2012-2013 tweets through visual inspection, topic modeling, and input from native Filipinos (Fig. 1). Topic modeling included Latent Dirichlet allocation (LDA) modeling using Gibbs sampling and n-grams methods along with keyword extraction and theme creation using PNNL-developed Rapid Automatic Keyword Extraction and Computational and Analysis of Significant Themes. The final lexicon was curated through a combination of these methods’ outputs and visual inspection of resulting tweets.

**Figure 1:**
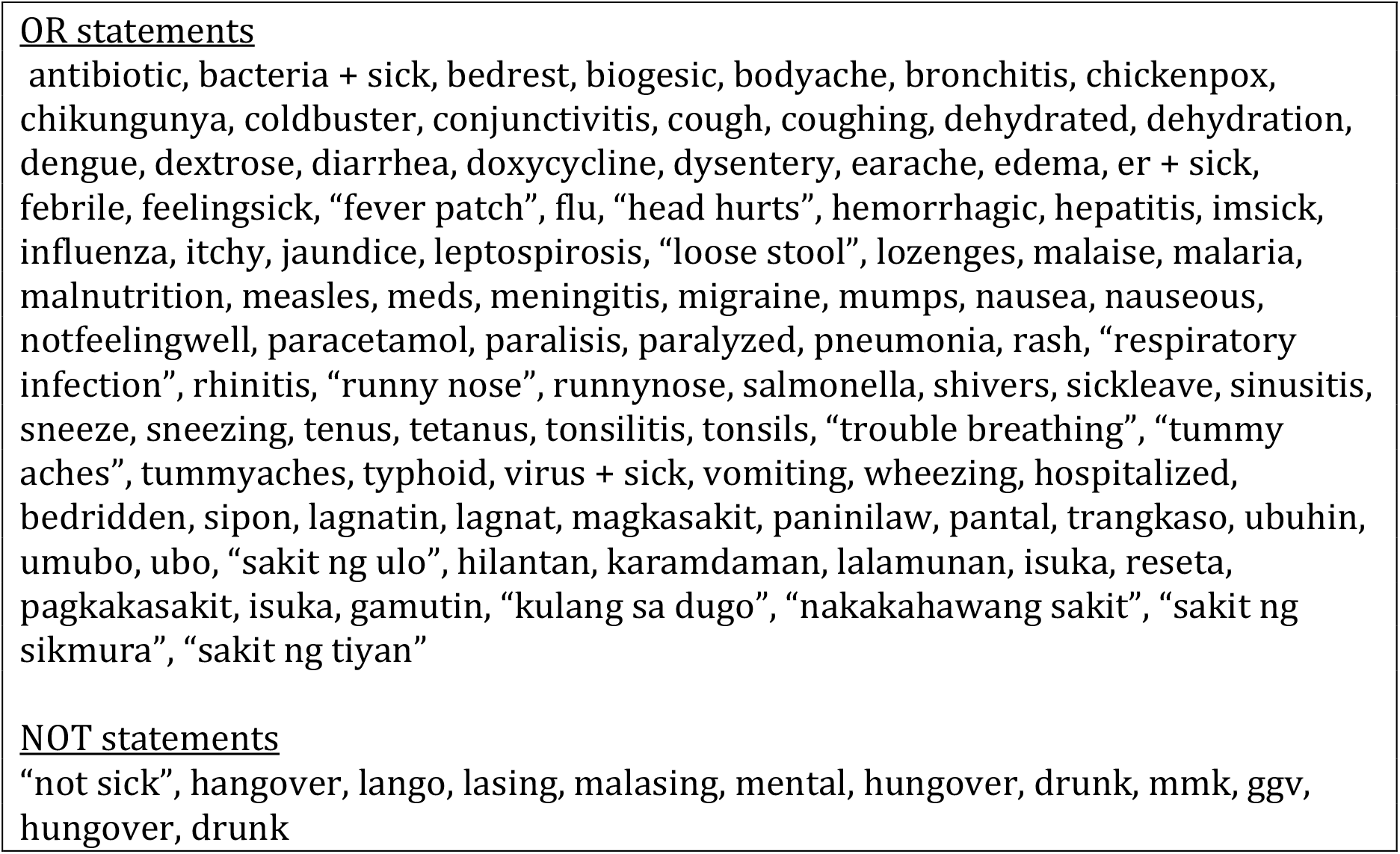
The lexicon of words and phrases used to filter out sick/disease related tweets. *Note that “+” means the words if they are in the same tweet regardless of order or spacing whereas “quoted words” need to be in the given order

### Outbreak Labels from Case Counts

To determine the reference parameter for disease outbreaks from weekly case count data, we ran a hidden markov model (HMM) for each of the diseases per region over 2012-2013. To identify the best-fit model, we ran mixed HMMs with two hidden states (R depmix library) for multiple families (i.e., Gaussian, Poisson, binomial, and multinomial) using the normal, log 10, and square root of the disease count data with parameters optimized (R Viterbi function) in each case. In general, cholera, leptospirosis, and measles were best modeled with a Poisson distribution, and dengue, ILI, and typhoid best fit a Gaussian distribution.

### Lexical-based Models

Time series data used in statistical models consisted of the number of tweets filtered by the disease lexicon by week and labeled with individual regions.

#### a. BreakoutDetection Model (Twitter R Program)

Twitter’s outbreak detection R program, BreakoutDetection, was used to identify time points in the data where there has been a significant change in the time series median indicative of an outbreak. Models and parameters were assessed by the weekly disease case counts per location. The program was run for individual island groups, regions, diseases, and diseases by region. The results are reported as positive and negative predictive values (PPV/NPV), sensitivity and specificity, and positive and negative likelihood ratios (PLR/NLR) of the outbreaks defined by BreakoutDetection compared to those predicted by HMM.

#### b. Autoregressive Integrated Moving Average with Explanatory Variable (ARIMAX) Model

An ARIMAX model was created for each disease by region using 2012 lexicon-filtered tweets and disease count data. A forecast ARIMAX model was run using the 2013 lexicon-filtered tweets and the 2012 model as its base. Accuracy measurements (i.e., mean error (ME), root mean squared error (RMSE), mean absolute error (MAE) and mean absolute scaled error (MASE)) of the forecasted model were calculated based on the actual 2013 disease counts for that disease-region pair. To be able to compare between datasets, the RMSE was normalized (NRMSE) to a percent by dividing by the difference between the maximum and minimum dependent values.

#### c. Regression Models

Multilinear regression (MLR) was used to model the disease case counts (dependent variable) using the number of lexicon-filtered tweets, province, region, year week, number of weeks from a disaster in that region, and disaster type. Depending on the distribution of the disease data, either a Poisson model or a generalized linear model was applied. Logistic Regression (LR) was used to model the HMM outbreak status (dependent variable) using the number of lexicon-filtered tweets, province, region, year week, number of weeks from a disaster in that region, and disaster type. For both MLR and LR, a model was created for all of the Philippines data for all diseases, all of the Philippines’ data for each disease individually, each island group by disease, and each region by disease. The best model was determined by the lowest Akaike information criterion (AIC) value, and predictors were considered significant for p-values <0.05. Note: Week here was treated as a number to proxy for the inherent time series correlation between weeks.

### Document Embedding Models (Fig. 2)

#### a. Disease State Model Building

Gensim’s Doc2Vec was used to create four embeddings consisting of tweet text grouped by location and labeled with tweet information (i.e., language and unique user), time of tweet (i.e., year week and month), location data (i.e., region and island group), regional disease data (i.e., disease type and week count, HMM outbreak state and corresponding disease type in outbreak), and regional disaster data (i.e., disaster types and weeks from disaster). Distributed memory (PV-DM) models were parameterized using various windows (distance between predicted word and context words) and size (dimensionality of the feature vectors). The workers were set to the maximum available and the minimum count to one (i.e., all words are important). By comparing model output to real data, the sliding window size was set to 10 and size of vector space to 100. The saved models were trained on all 2012 and 2013 data through random shuffling of 10 epochs.

**Figure 2:**
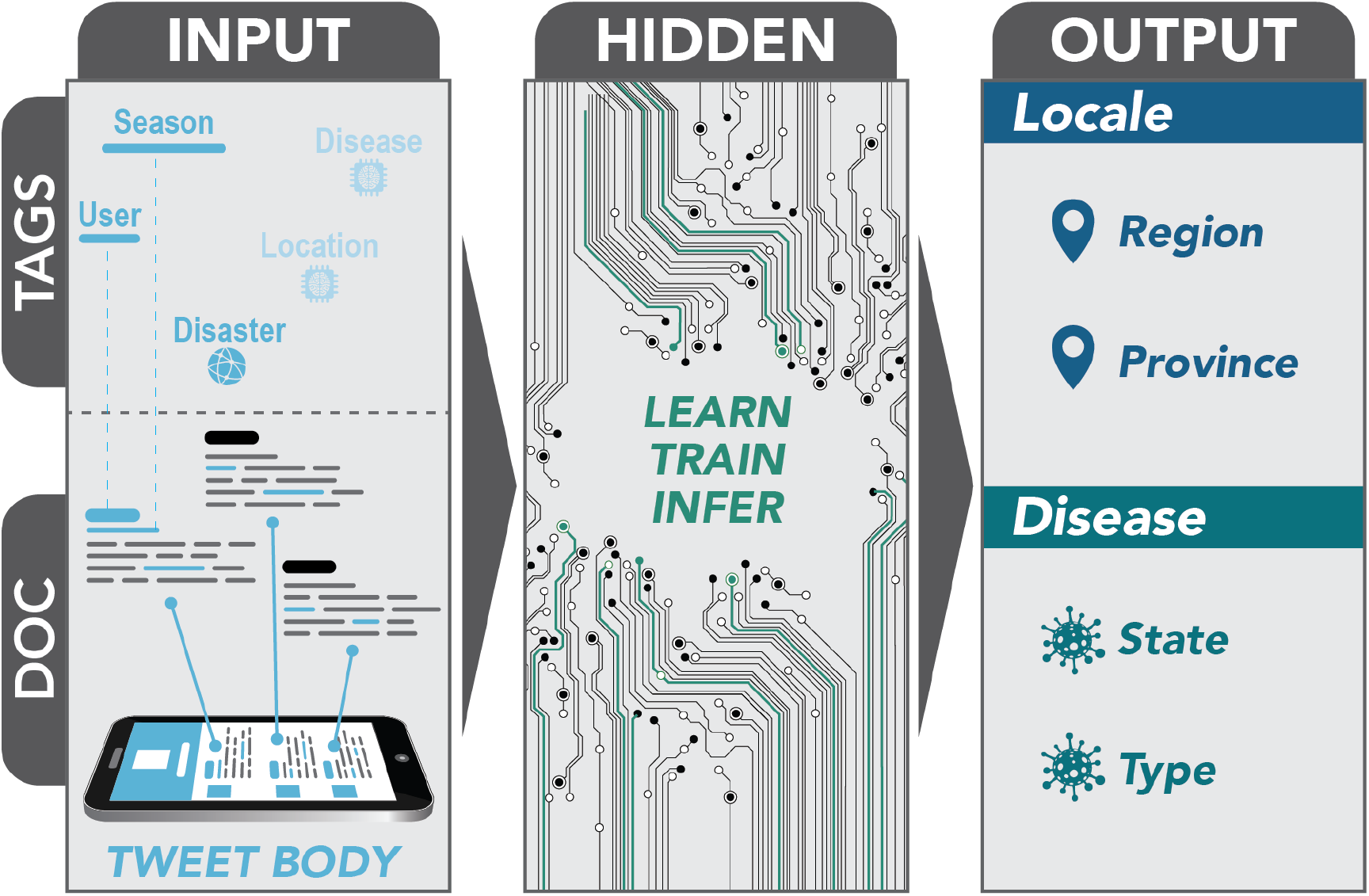
The neural network framework developed to identify the location and disease state of the location based on a tweet body, user, date, and weeks from a disaster type. The network was trained on this information plus information on disease type, case counts, and regional location.

#### b. Disease State Model Analysis

The accuracy of the individual document embedding models were tested against all 2012–2013 tweets by day and summarized by week. The method was designed to test the implementation of the network to geo-tagged tweets collected daily. However, to compare to the weekly disease data, the results were summarized by week. The final evaluation examined PPV, NPV, sensitivity, specificity, PLR, and NLR of the models by island group, region, disease type, and disease by region compared to the HMM outbreak status (similar to the BreakoutDetection results) over the full two years of data.

To produce these results, the tweets were converted into Doc2Vec format containing the tweet text and tagged with tweet information (language and unique user), time of tweet (month, year week, and region_day), location data (region and island group), and disaster information (type and weeks from disaster; optional). Then, each tweet was vectorized based on the network model for the same location. The cosine similarity value (i.e., the cosine between two vectors) was computed for the individual tweet vector and specific vectors within the larger network model, i.e., those that are labeled with disease outbreak, no outbreak, and the type of disease. A positive cosine similarity value (0-1) indicates that the vectors are similar, whereas a negative value implies the vectors are different.

## Results

### Tweets

Between 2012 and 2013, 50 million tweets geo-tagged were collected from the Philippines. There were about 80,700 tweets across the 17 regions (0.1% - 0.2% of total tweets per region) filtered by the disease lexicon (Table 1). The total number of tweets and, subsequently, the lexicon-filtered tweets varied highly by location and ranged from 50,565 in the Autonomous Region in Muslim Mindanao (ARMM) to 22,725,791 in the National Capitol Region (NCR).

### Diseases

Through the HMM models, the disease outbreaks by region varied by location and disease (Table 2). Typhoid, dengue, and ILI outbreaks occurred most frequently throughout the time period and regions V, VI, and NCR (Fig. 3) [15]. Summary statistics for the case counts per region are shown in Table 3.

**Table 2:** Number of weeks a disease outbreak was predicted by HMM in a given region over the 2-year period (104 weeks) between 2012-2013 in the Philippines.

| Island Group | Region | CHOL | DEN | ILI | LEPT | MEAS | TY | Total |
| --- | --- | --- | --- | --- | --- | --- | --- | --- |
| Luzon | NCR | 3 | 41 | 61 | 35 | 13 | 83 | 236 |
|  | CAR | 0 | 47 | 35 | 21 | 24 | 71 | 198 |
|  | I | 0 | 52 | 37 | 23 | 0 | 63 | 175 |
|  | II | 0 | 35 | 46 | 40 | 4 | 71 | 196 |
|  | III | 1 | 64 | 43 | 30 | 9 | 41 | 188 |
|  | IVA | 12 | 49 | 37 | 9 | 16 | 67 | 190 |
|  | IVB | 10 | 16 | 74 | 0 | 8 | 32 | 140 |
|  | V | 39 | 44 | 12 | 71 | 1 | 72 | 239 |
| Visayas | VI | 19 | 31 | 40 | 13 | 70 | 72 | 245 |
|  | VII | 24 | 60 | 61 | 4 | 17 | 4 | 170 |
|  | VIII | 5 | 53 | 47 | 50 | 6 | 58 | 219 |
| Mindanao | IX | 4 | 45 | 64 | 18 | 8 | 87 | 226 |
|  | X | 37 | 12 | 16 | 4 | 0 | 58 | 127 |
|  | XI | 0 | 47 | 46 | 18 | 1 | 1 | 113 |
|  | XII | 1 | 37 | 48 | 16 | 3 | 66 | 171 |
|  | XIII | 67 | 77 | 25 | 6 | 1 | 29 | 205 |
|  | ARMM | 1 | 29 | 42 | 2 | 0 | 87 | 161 |
| Total |  | 223 | 739 | 734 | 360 | 181 | 962 |  |

**Table 3.**
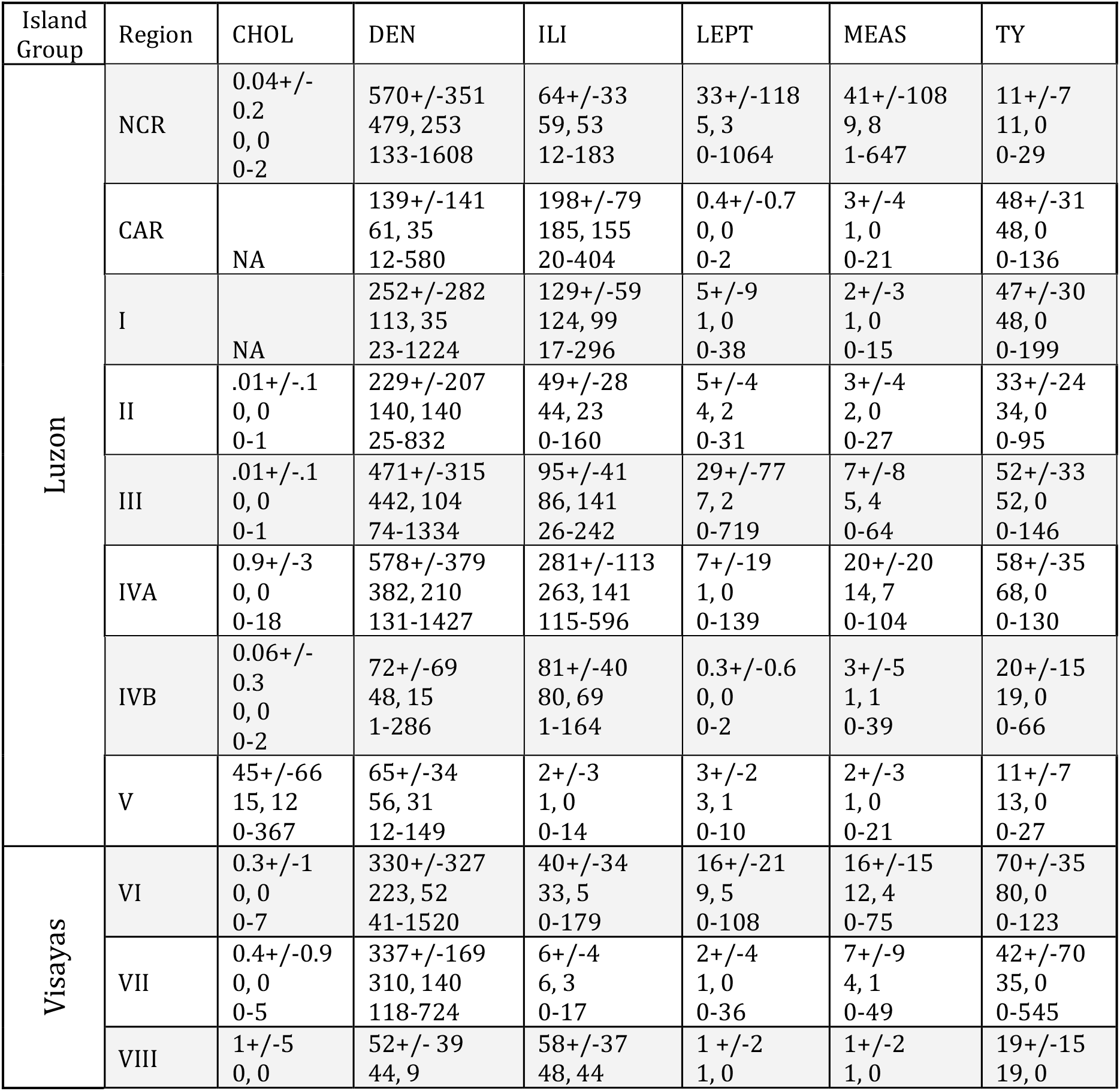
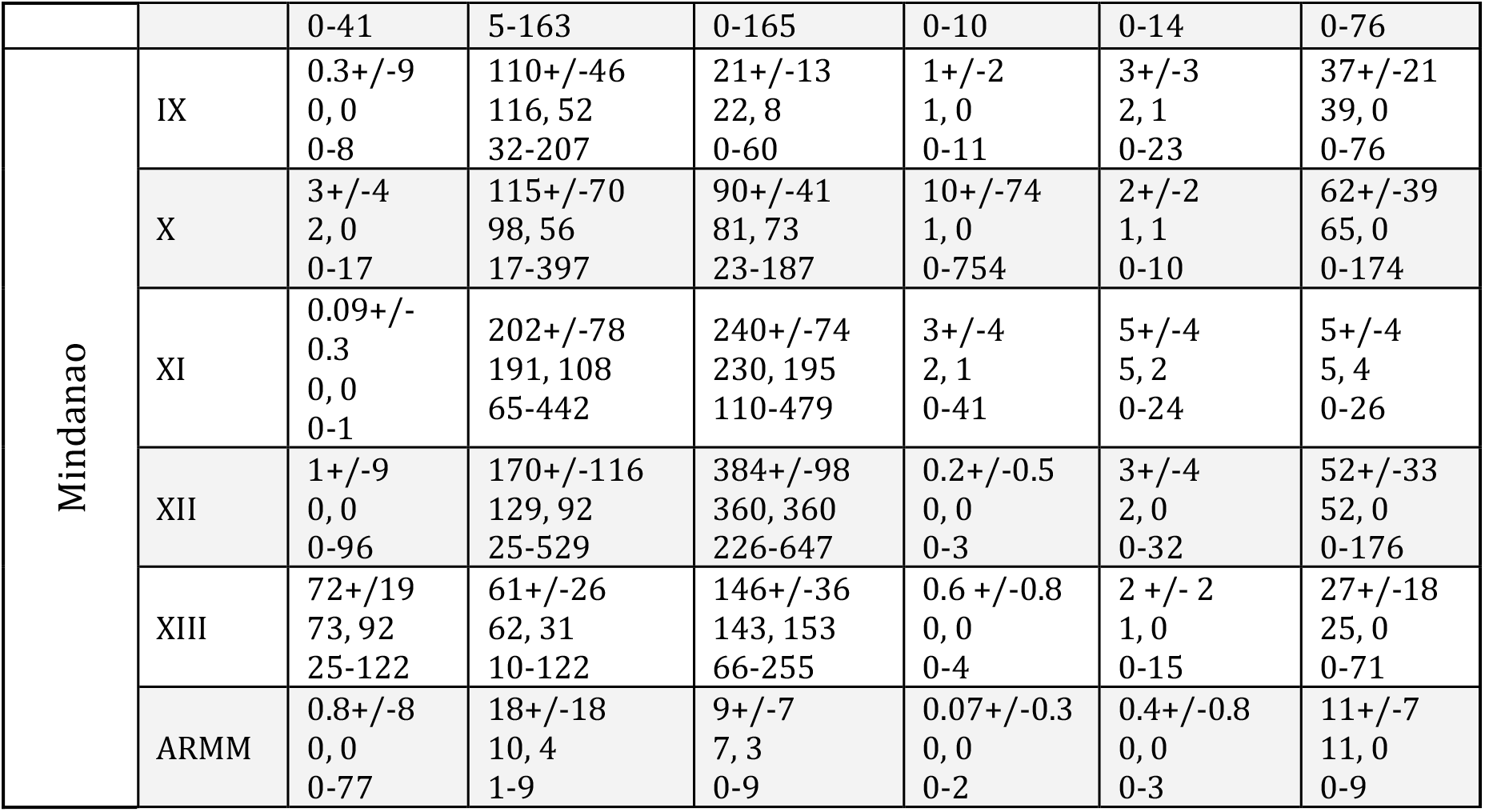
Mean +/-standard deviation (row 1), median, mode (row 2), range of cases between 2012 and 2013 for the six diseases over the all regions.

| Island Group | Region | CHOL | DEN | ILI | LEPT | MEAS | TY |
| --- | --- | --- | --- | --- | --- | --- | --- |
| Luzon | NCR | 0.04+/-0.2<br>0, 0<br>0-2 | 570+/-351<br>479, 253<br>133-1608 | 64+/-33<br>59, 53<br>12-183 | 33+/-118<br>5, 3<br>0-1064 | 41+/-108<br>9, 8<br>1-647 | 11+/-7<br>11, 0<br>0-29 |
|  | CAR | NA | 139+/-141<br>61, 35<br>12-580 | 198+/-79<br>185, 155<br>20-404 | 0.4+/-0.7<br>0, 0<br>0-2 | 3+/-4<br>1, 0<br>0-21 | 48+/-31<br>48, 0<br>0-136 |
|  | I | NA | 252+/-282<br>113, 35<br>23-1224 | 129+/-59<br>124, 99<br>17-296 | 5+/-9<br>1, 0<br>0-38 | 2+/-3<br>1, 0<br>0-15 | 47+/-30<br>48, 0<br>0-199 |
|  | II | .01+/-0.1<br>0, 0<br>0-1 | 229+/-207<br>140, 140<br>25-832 | 49+/-28<br>44, 23<br>0-160 | 5+/-4<br>4, 2<br>0-31 | 3+/-4<br>2, 0<br>0-27 | 33+/-24<br>34, 0<br>0-95 |
|  | III | .01+/-0.1<br>0, 0<br>0-1 | 471+/-315<br>442, 104<br>74-1334 | 95+/-41<br>86, 141<br>26-242 | 29+/-77<br>7, 2<br>0-719 | 7+/-8<br>5, 4<br>0-64 | 52+/-33<br>52, 0<br>0-146 |
|  | IVA | 0.9+/-3<br>0, 0<br>0-18 | 578+/-379<br>382, 210<br>131-1427 | 281+/-113<br>263, 141<br>115-596 | 7+/-19<br>1, 0<br>0-139 | 20+/-20<br>14, 7<br>0-104 | 58+/-35<br>68, 0<br>0-130 |
|  | IVB | 0.06+/-0.3<br>0, 0<br>0-2 | 72+/-69<br>48, 15<br>1-286 | 81+/-40<br>80, 69<br>1-164 | 0.3+/-0.6<br>0, 0<br>0-2 | 3+/-5<br>1, 1<br>0-39 | 20+/-15<br>19, 0<br>0-66 |
|  | V | 45+/-66<br>15, 12<br>0-367 | 65+/-34<br>56, 31<br>12-149 | 2+/-3<br>1, 0<br>0-14 | 3+/-2<br>3, 1<br>0-10 | 2+/-3<br>1, 0<br>0-21 | 11+/-7<br>13, 0<br>0-27 |
| Visayas | VI | 0.3+/-1<br>0, 0<br>0-7 | 330+/-327<br>223, 52<br>41-1520 | 40+/-34<br>33, 5<br>0-179 | 16+/-21<br>9, 5<br>0-108 | 16+/-15<br>12, 4<br>0-75 | 70+/-35<br>80, 0<br>0-123 |
|  | VII | 0.4+/-0.9<br>0, 0<br>0-5 | 337+/-169<br>310, 140<br>118-724 | 6+/-4<br>6, 3<br>0-17 | 2+/-4<br>1, 0<br>0-36 | 7+/-9<br>4, 1<br>0-49 | 42+/-70<br>35, 0<br>0-545 |
|  | VIII | 1+/-5<br>0, 0 | 52+/-39<br>44, 9 | 58+/-37<br>48, 44 | 1+/-2<br>1, 0 | 1+/-2<br>1, 0 | 19+/-15<br>19, 0 |
|  |  | 0-41 | 5-163 | 0-165 | 0-10 | 0-14 | 0-76 |
| Mindanao | IX | 0.3+/-9<br>0, 0<br>0-8 | 110+/-46<br>116, 52<br>32-207 | 21+/-13<br>22, 8<br>0-60 | 1+/-2<br>1, 0<br>0-11 | 3+/-3<br>2, 1<br>0-23 | 37+/-21<br>39, 0<br>0-76 |
|  | X | 3+/-4<br>2, 0<br>0-17 | 115+/-70<br>98, 56<br>17-397 | 90+/-41<br>81, 73<br>23-187 | 10+/-74<br>1, 0<br>0-754 | 2+/-2<br>1, 1<br>0-10 | 62+/-39<br>65, 0<br>0-174 |
|  | XI | 0.09+/-<br>0.3<br>0, 0<br>0-1 | 202+/-78<br>191, 108<br>65-442 | 240+/-74<br>230, 195<br>110-479 | 3+/-4<br>2, 1<br>0-41 | 5+/-4<br>5, 2<br>0-24 | 5+/-4<br>5, 4<br>0-26 |
|  | XII | 1+/-9<br>0, 0<br>0-96 | 170+/-116<br>129, 92<br>25-529 | 384+/-98<br>360, 360<br>226-647 | 0.2+/-0.5<br>0, 0<br>0-3 | 3+/-4<br>2, 0<br>0-32 | 52+/-33<br>52, 0<br>0-176 |
|  | XIII | 72+/-19<br>73, 92<br>25-122 | 61+/-26<br>62, 31<br>10-122 | 146+/-36<br>143, 153<br>66-255 | 0.6 +/-0.8<br>0, 0<br>0-4 | 2 +/- 2<br>1, 0<br>0-15 | 27+/-18<br>25, 0<br>0-71 |
|  | ARMM | 0.8+/-8<br>0, 0<br>0-77 | 18+/-18<br>10, 4<br>1-9 | 9+/-7<br>7, 3<br>0-9 | 0.07+/-0.3<br>0, 0<br>0-2 | 0.4+/-0.8<br>0, 0<br>0-3 | 11+/-7<br>11, 0<br>0-9 |

**Figure 3.**
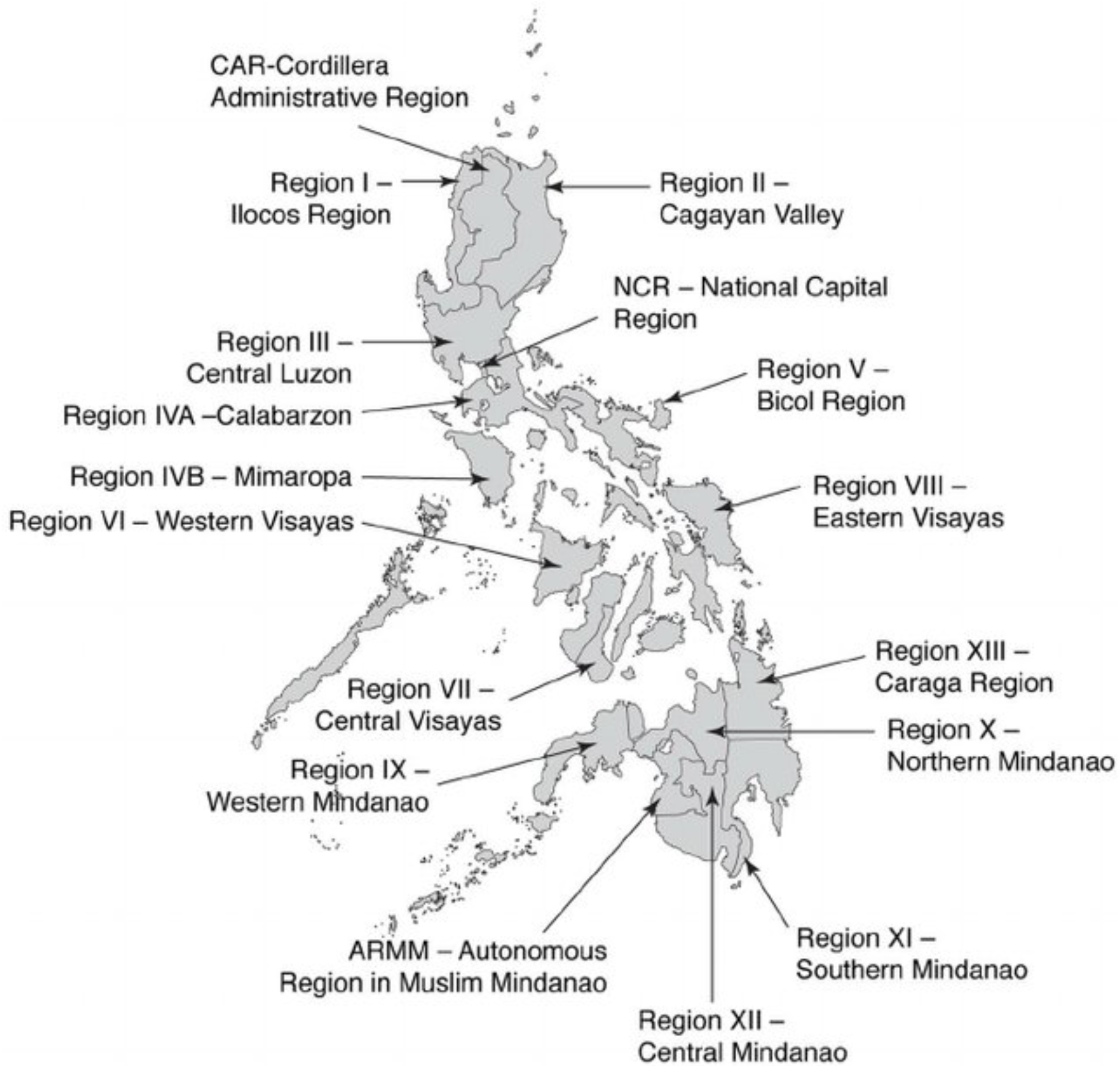
Map of the administrative regions of the Philippines. Image reprinted from [15].

### Disasters

During 2012–2013, there were instances of tropical cyclones (n = 16; n = 29), flooding (n = 26; n = 23), landslides (n = 5; n = 12), earthquakes of significant magnitude (n = 9; n = 9), tornados (n = 3; n = 7), and volcanic explosions (n = 1; n = 1).

### BreakoutDetection Model

Overall, the models had a high negative predictive value (mean = 0.70 +/-0.20) and a low positive predictive value (mean = 0.33 +/- .31). Along the same lines, the models had a high specificity (mean = 0.95 +/-0.03) but a low sensitivity (mean = 0.06 +/-0.06). The overall PLR was 1.86 +/-3.48 with NLR of 0.99 +/-0.07.

In terms of individual models, the region III had the best overall results with highlighted models for ILI and typhoid. The NCR region was very similar in overall results with the top disease models for that region being for cholera, ILI, and typhoid. Region V was also comparable with specific top disease models for cholera, leptospirosis, and typhoid. The last mentionable region is IVB with a top cholera model. Other region-specific disease models that had a high positive likelihood value, low negative likelihood value, and high specificity were region VII cholera, region XI for leptospirosis, and region VI for ILI.

Models for ILI in regions II, III, VI, and cholera in region VII had greater than 0.67 PPV/NPV, specificity greater than 0.97, and PLR between 4 and 22. However, sensitivity was between 0.12 and 0.21 with an average NLR of 0.85. Other models are not worth noting here.

### Autoregressive Integrated Moving Average with Explanatory Variable Model

As a measure of over-fitness, the difference between the training and test sets NRMSE for all models has a median of 0.42% with a minimum value of 0.01% and maximum value of 4956%. In general, 93% of the models performed very well with an NRMSE of < 3.0% (95/102); however, predictive models of cholera for NCR, II, IVA, VI, VII, and XI and typhoid for NCR over-fit the data and produced large differences in errors between the training and testing data (NRMSE range 3% to 4955%). For cholera, regions CAR, I, and III were not modeled due to lack of data. Using the MASE as the measurement of error because it can also be interpreted across various types of grouped time series models, the models (region-disease) that had a MASE of one or less are ARMM-LEPTO, IVB-CHOL, and ARMM-MEAS.

### Logistic Regression

Logistic regression (LR) was used to model the HMM outbreak state based on tweets, location (province and region), week, and number of weeks from last disaster and type. There were 16 LR models those top AIC value contained significant tweets (Table 4). There were four main results seen in these models: (1) dengue was the best modeled by tweets; (2) two models (dengue) included disasters as an important predictive variable; (3) eight of these models contained weeks and tweets; and (4) five models were based on region location and tweets only.

**Table 4.** Top logistic regression models for disease outbreak (dependent variable) based on HMM outbreak models and corresponding independent variables based on stepwise AIC selection and include tweet counts as a significant variable. The “--” cells mean that the variable was not in the model; regions labeled with “Multiple” mean that there were contained multiple significant regions (n); bolded cells describe the filter for the dependent variable.

| Disease | Province | Region | Week<br>(p-value) | Disaster (type; #<br>weeks \| p-value) | Tweets<br>(p-value) | AIC |
| --- | --- | --- | --- | --- | --- | --- |
| **CHOL** | **Visayas** | VII, VIII | -- | -- | 4.96E-06 | 231.7 |
| **DEN** | -- | Multiple (n=12) | 4.51E-04 | Flood; 0-5 weeks | 3.90E-02 | 2082.3 |
|  | **Luzon** | III, IVA, IVB, V | 1.72E-06 | Storm; 0-5 weeks | 2.58E-02 | 939.1 |
|  |  | **IVA** | > 0.05 | > 0.05 | 2.67E-02 | 123.9 |
|  | **Mindanao** | IX, XI, XIII | 5.56E-06 | -- | 3.14E-03 | 731.3 |
|  |  | **IX** | 1.55E-02 | -- | 2.12E-02 | 140.8 |
|  |  | **XII** | 4.12E-07 | -- | 1.00E-02 | 98.6 |
|  | **Visayas** | **VIII** | -- | -- | 2.94E-03 | 123.9 |
| **ILI** | -- | Multiple (n=7) | -- | -- | 1.61E-05 | 2231.1 |
|  | **Luzon** | NCR, III, IVA/B, V | -- | -- | 3.39E-05 | 1023.2 |
|  |  | **II** | > 0.05 | -- | 2.67E-02 | 128.6 |
| **LEPT** | **Luzon** | II, IVA, V | > 0.05 | -- | 8.92E-04 | 809.7 |
|  |  | **II** | 1.08E-04 | -- | 1.61E-03 | 125.3 |
| **MEAS** | **Visayas** | VII, VIII | -- | -- | 2.98E-02 | 273 |
| TY | Luzon | IVB | 4.04E-06 | -- | 2.92E-02 | 100.5 |
|  | Mindanao | XII | 8.03E-07 | -- | 1.34E-02 | 104.5 |

### Multilinear Regression

Multilinear regression was used to model the disease case counts through various filters of disease type, province, and region with predictors of lexicon tweet counts, week of year, disaster type, and weeks from last disaster. Forty-eight out of the 102 top models (47%) contained tweet counts as a significant variable (Table 5). Of these models, disasters were significant in 40% (19/48) and weeks were significant in 79% (38/48). There were no common themes between models and the disaster types, weeks from last disaster, or direction of correlation. All three island groups were represented in each type of disease model including all diseases combined, except Visayas did not have a tweet significant model for ILI and TY. AIC values were fairly high for models based on all of the Philippines or a full island group (mean = 16,691; range = 777 – 134,898) but for region-specific models, the AIC is lower (mean = 989; range = 331-3953). Specific regions that were best represented by these models are Luzon-IVA, Luzon-NCR, Visayas-VIII, and Mindanao-all.

**Table 5.** Top MLR models for disease counts (dependent variable) and corresponding independent variables based on stepwise AIC selection and include tweet counts as a significant variable. The “--” cells mean that the variable was not in the model; regions labeled with “Multiple” mean that there were contained multiple significant regions (n); bolded cells describe the filter for the dependent variable; for models using Poisson distribution, only AIC is reported.

| Dis - ease | Province | Region | Week (p-value) | Disaster (type; weeks \| p-value) | Tweets (p-value) | Adj R<sup>2</sup> | AIC |
| --- | --- | --- | --- | --- | --- | --- | --- |
| ALL | -- | Multiple (n=14) | 1.94E-02 | -- | 1.43E-02 | 0.07 | 134898 |
| CHOL | Luzon | NCR, I, II, IVA, V | 2.48E-05 | Flood; not 0-2 | 1.56E-05 | 0.91 | 9154 |
|  | Visayas | VIII | < 2.00E-16 | Random | < 2.00E-16 | -- | 777 |
|  |  | VIII | 2.19E-04 | -- | 4.61E-03 | 0.12 | 620 |
|  | Mindanao | X | -- | 4 types; 0-4, 6, 10 | 1.74E-11 | -- | 442 |
|  |  | XII | < 2.00E-16 | -- | < 2.00E-16 | -- | 603 |
|  |  | XIII | 1.52E-08 | -- | 3.10E-03 | -- | 1100 |
|  |  | ARMM | 1.11E-10 | -- | 1.02E-07 | -- | 611 |
| DEN | Luzon | IVA | -- | > 0.05 | 4.82E-02 | 0.41 | 1509 |
|  | Visayas | VIII | -- | -- | 1.08E-05 | 0.32 | 4223 |
|  |  | VI | -- | -- | 1.49E-04 | 0.12 | 1490 |
|  |  | VIII | > 0.05 | -- | 3.87E-03 | 0.19 | 1041 |
|  | Mindanao | IX - XIII | 1.48E-08 | -- | 1.77E-03 | 0.48 | 7019 |
|  |  | IX | > 0.05 | -- | 1.01E-02 | 0.05 | 1092 |
|  |  | XII | 8.57E-14 | -- | 4.07E-04 | 0.21 | 956 |
|  |  | XIII | 1.54E-03 | -- | 2.84E-02 | 0.2 | 955 |
| ILI | -- | Multiple (n=13) | 4.15E-03 | -- | 2.47E-03 | 0.79 | 19109 |
|  | Luzon | NCR, I - V | -- | -- | 3.09E-02 | 0.67 | 9154 |
|  |  | NCR | 1.66E-14 | -- | 3.32E-03 | 0.6 | 932 |
|  |  | IVA | 1.08E-04 | -- | 1.19E-02 | 0.14 | 1267 |
|  | Mindanao | X - XIII | 4.50E-05 | -- | 1.26E-03 | 0.86 | 6735 |
|  |  | XI | -- | -- | 9.49E-05 | 0.13 | 1180 |
|  |  | ARMM | 3.18E-04 | full range | 4.01E-02 | 0.77 | 589 |
| LEPT | -- | Multiple (n=16) | < 2.00E-16 | full range | < 2.00E-16 | -- | 26483 |
|  | Luzon | NCR, I - V | 2.15E-11 | full range | < 2.00E-16 | -- | 12428 |
|  |  | **NCR** | > 0.05 | full range | 1.90E-09 | -- | 3953 |
|  |  | **I** | 2.76E-02 | -- | 2.23E-02 | -- | 1343 |
|  |  | **II** | 8.74E-11 | -- | 2.52E-04 | -- | 633 |
|  |  | **III** | < 2.00E-16 | full range | < 2.00E-16 | -- | 1189 |
|  |  | **IVA** | < 2.00E-16 | 5 types; 0-5, 18+ | < 2.00E-16 | -- | 974 |
|  | **Visayas** | VII, VIII | -- | Flood; 1-3, 7-14 | 6.28E-03 | 0.79 | 2138 |
|  |  | **VIII** | 3.69E-02 | -- | 4.14E-02 | -- | 331 |
|  | **Mindanao** | IX, X, XI, XIII | 3.53E-09 | 4 types; 0-3, 17 | < 2.00E-16 | -- | 3987 |
|  |  | **X** | < 2.00E-16 | 6 types; 0-8, 11+ | < 2.00E-16 | -- | 829 |
| **MEAS** | -- | NCR, III, IVA, VI, VII | 1.91E-10 | -- | < 2.00E-16 | 0.54 | 15579 |
|  | **Luzon** | NCR, III, IVA | 5.82E-06 | -- | < 2.00E-17 | 0.55 | 7902 |
|  |  | **NCR** | 1.35E-02 | 3 types | 2.86E-12 | 0.75 | 1160 |
|  |  | **III** | -- | 5 types | 1.05E-11 | -- | 547 |
|  |  | **IVA** | -- | Storm; 32-33 | 1.23E-14 | 0.74 | 816 |
|  | **Visayas** | VIII | 1.42E-02 | full range | 7.34E-11 | -- | 2630 |
|  |  | **VI** | < 2.00E-16 | full range | 3.08E-10 | -- | 1038 |
|  |  | **VIII** | 9.67E-06 | -- | 1.95E-03 | 0.16 | 464 |
|  | **Mindanao** | IX - XIII | -- | -- | 2.60E-02 | 0.22 | 3102 |
|  |  | **XI** | 3.65E-10 | None, 19-20 | 1.66E-03 | -- | 572 |
| **TY** | **Luzon** | **NCR** | 5.38E-08 | -- | 1.40E-04 | 0.26 | 671 |
|  |  | **IVB** | 3.08E-02 | -- | 8.34E-03 | 0.05 | 853 |
|  | **Mindanao** | IX - XIII | 1.19E-04 | -- | 3.12E-02 | 0.43 | 5727 |
|  |  | **IX** | 4.99E-07 | -- | 1.85E-02 | 0.23 | 905 |

### Document Embeddings

The neural network was modeled on all 2012–2013 data and then tested on individual days combined into weeks for the same time period. These results were compared to HMM outbreak results derived from case counts (Table 6). For the comparison of data on the island group level, Luzon was the only region that predicted any positive results although the sensitivity of the model was only 0.01 (PPV = 0.13; PLR = 0.31) with a specificity of 0.96 (NPV = 0.66; NLR = 1.03). On the region level, only Luzon’s CAR and NCR predicted any positive results both had low PPV (0.22/0.13), sensitivity (0.01/0.03) and PLR (0.60/0.23) with high specificity (0.98/0.88), respectively.

**Table 6.**
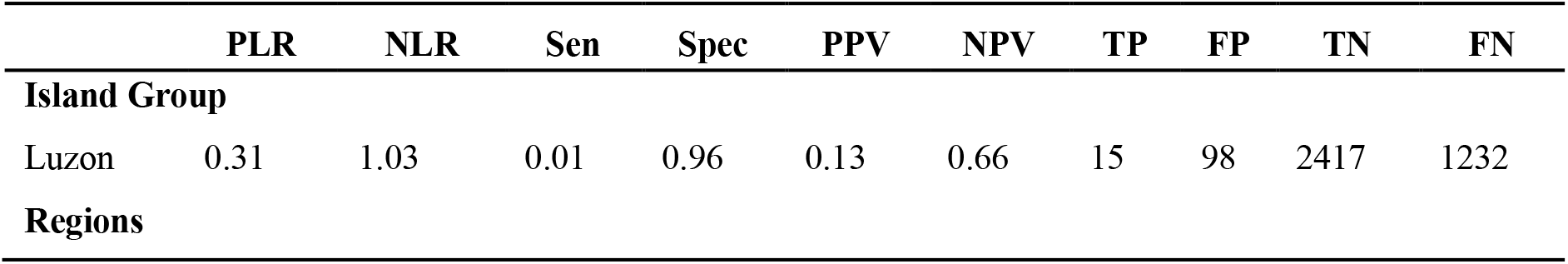
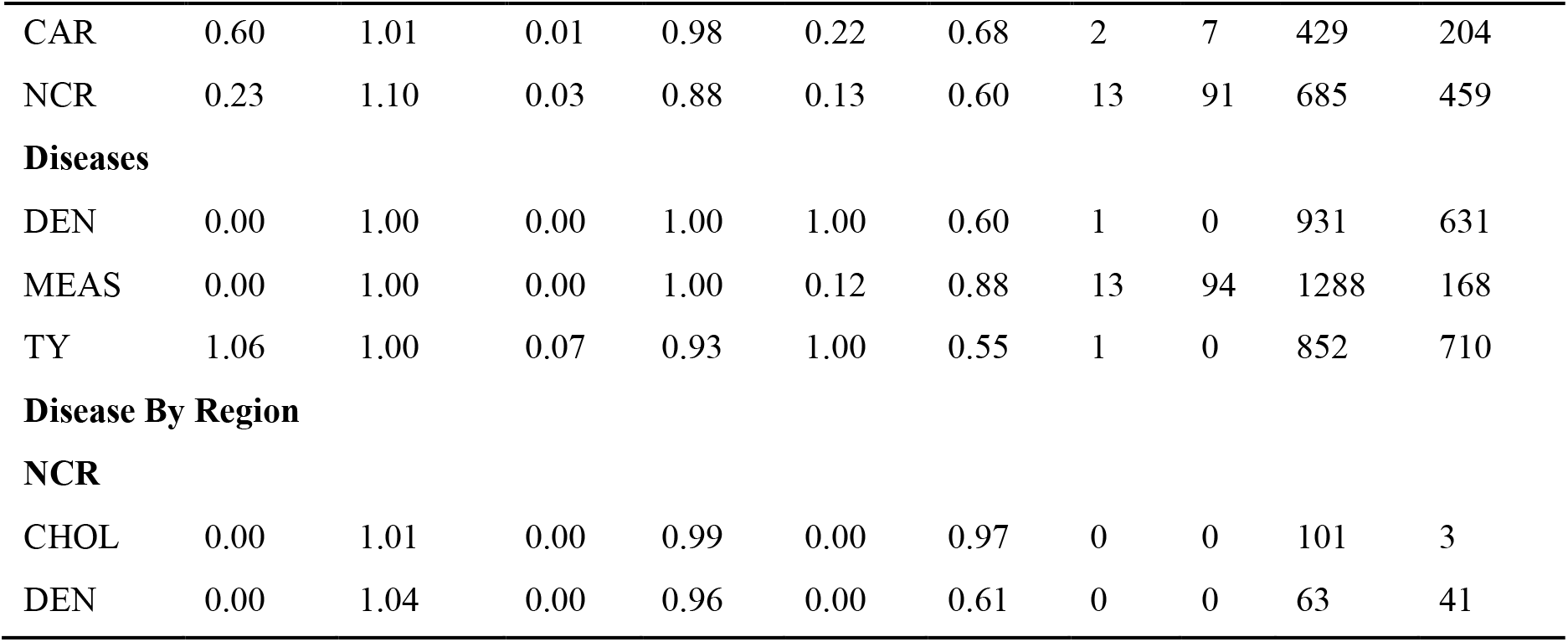
Doc2Vec results from 2012-2013 data tested against HMM predicted outbreaks from disease case counts. Only results that contained both positive and negative outbreak results to use for calculating accuracy measurements are shown. Positive likelihood ratio (PLR) = =Sen/(1-Spec); Negative likelihood ratio (NLR) = =(1-sen)/spec; Sensitivity (Sen) = =TP/(TP+FN); Specificity (Spec) = =TN/(FP+TN); Positive predictive value (PPV) = TP/(TP+FP); Negative predictive value (NPV) = TN/(TN+FN); True positive (TP); False positive (FP); True negative (TN); False negative (FN).

## Discussion

The goal of this research was to identify the potential disease outbreaks post-natural disaster recovery in the Philippines with only the use of Twitter data and type/time since last disaster. This research was complicated by the varying language usage in tweets (English, Tagalog, Taglish, and more than 100 native dialects), Twitter use in general (e.g., NCR had 450 times the number of tweets than ARMM), and the high number of disasters present (141 total). All considered, the analyses show that the use of only tweets to predict disease outbreaks in the Philippines has varying results that depend on which technique is being applied, the disease type, and location discussed below. However, the number of tweets used in this analysis from 2012 and 2013 seems to be doubling in the following years and, therefore, consistency in results may increase in future data analyses.

### Data

Twitter data, in general, has been shown to be very noisy when trying to extract a specific topic, ex. Influenza [16], which can be cofounded overtime by new arising topics [17]. Due to these known issues, we focused our lexicon-based analyses on general disease state or not. We only attempted to identify specific infectious disease topics when using machine learning document embedded models trained on all available tweets (not lexicon-based).

Disease count data, when captured in real time, can cause issues because of the amount of time it takes to report a disease [18]. To avoid this problem, we only used historical disease data to help understand the disease trends in the underlying population. However, an underlying issue that we encountered was that we did not have the ground truth on outbreak status. We had disease case counts by week and used HMM models to determine the historical outbreak status of the region for a specific disease. This method enabled the most flexibility in algorithm choice for assessing the best method for increased levels of disease in a given geographic area.

### Modeling Disease Outbreaks

For those models that detected the presence or absence of an outbreak (i.e., BreakoutDetection and Doc2Vec) based on the HMM-derived outbreaks, the validity of the results is best represented by likelihood ratios since the true prevalence and outbreak status of the disease is unknown. A likelihood ratio of greater than one indicates the test result is associated with the disease. A likelihood ratio less than one indicates that the result is associated with absence of the disease. Tests where the likelihood ratios lie close to one have little practical significance. Other accuracy measurements used are the PPV and NPV (i.e., the proportions of positive and negative results that are true positive and true negative results, respectively) and sensitivity and specificity (i.e., true positive and true negative rates, respectively).

BreakoutDetection, which is a lexicon-derived time series model used to predict outbreaks, was generally good at negative predictions of outbreaks but not as good at positive predictions (i.e., high NPV and specificity but low PPV and sensitivity). These conclusions are similar to the results from Gensim’s Doc2Vec, a neural network model also used to predict outbreaks. However, for BreakoutDetection, there were some very good predictive models for each island group, more specifically in seven out of the 17 regions (NCR, III, IVB, V, VI, VII, and XI) for cholera (n = 4), ILI (n = 3), typhoid (n = 3), and leptospirosis (n = 2). Gensim’s Doc2Vec only identified positive outbreaks in two regions (NCR and CAR) within one island group and the models still contained low PLR, PPV, and sensitivity with high specificity. The poor results for the neural network are most likely due to the complexity in language and diversity in topics in the Philippines Twitter dataset. Although this study contains a much larger dataset than other studies that used neural networks to classify disease status with text data [19-21], Twitter data is an indirect indicator of health status and therefore, signals are more hidden than traditional diagnostic and medical report data. Here, the neural network is unable to find a disease outbreak signal in the tweets with their inherent complexity coupled with the reduction in dataset dimensionality created by subsetting tweets by location and disease classification by weekly reports. By increasing the time period of the training set, i.e., more tweets and disease weeks, the neural network should perform better.

To complement the models above, which attempted to predict disease outbreaks, LR was used to model the relationship between an outbreak and potential predictive characteristics (i.e., lexicon-filtered tweets, location, week, and number of weeks from the last disaster in the region and type). Out of the 102 total LR models, only 16 contained tweets as a significant factor in disease outbreak status. Out of these models, there were eight region-specific (i.e., II [n = 2], IVA, IVB, VIII, IX, and XII [n = 2]), six island group-specific (i.e., Luzon [n=3], Mindanao, and Visayas [n=2]) and two disease-specific (i.e., dengue and ILI) models. Note: dengue and ILI were the only diseases that had multiple outbreaks per year in every region and island group (Table 2). Also, only two of these tweet-significant models identified disaster type as another significant factor and, therefore, may not play a large role in disease outbreak prediction when using lexicon-filtered tweets.

### Modeling Disease Case Counts

For comparison, the empirical disease case counts were directly used in both time series models (ARIMAX) and regression models (MLR). These models seemed to perform better than those using outbreaks as the dependent variable. This may be due to the discrete versus continuous nature of the variables or the statistical methods used to model the data.

For the 2012 date, disease case counts and filtered tweets were used to train the ARIMAX time series model and then only filtered tweets from 2013 were used to test it. These models performed extremely well except for cholera (9/17 regions) and typhoid (1 region), which is the opposite of the outbreak time series findings with BreakoutDetection where cholera and typhoid had the best predictive models.

Multilinear regression on case counts seemed to perform better than LR models using HMM outbreak status, similar to Culotta [22] and Bodnar and Salathé [23]. Here, 47% of the models incorporated filtered tweets as a significant factor. Disaster seemed to play a larger role as a correlation factor for case counts than outbreaks. However, the actual specific types of disasters, weeks from disaster, and direction of correlation varied widely causing the interpretation of these results to be confusing. Of these models, all island groups and almost all regions were represented in a region-disease specific model except region CAR, V, and VII. In addition, at the country level for all diseases, the top model contained tweets and weeks both at a p-value < 0.05.

## Conclusion

This study aimed to assess the utility of social media for identification of potential disease outbreak situations in the Philippines after natural disaster events. We have shown that publicly available Twitter data in various location-specific disease pairs is a significant factor in disease prediction. Our results also show that the time period since the last natural disaster may also play a role. However, there is high variability in the number of tweets by location and the types of diseases present, which is most likely affecting the predictive power of the tweets and time to disaster. In this case, the dataset should be split by disease type and tweet geolocation for better predictions.

We have compared various mathematical methods (e.g., statistical time series to machine learning neural networks) in effort to identify the algorithm with the best prediction capability. Based on our data, the “best” method varies by location and disease type. The neural network should have identified the location, disaster, and Twitter-specific attributes but most likely due to the minimal size and complexity of the dataset, our results were not as accurate as we expected. In terms of time series statistical models, the ARIMAX model outperformed the other models in most instances. We want to note that the input of disease case counts showed a promising improvement to the predictive capability of the model, i.e., the farther away from the trained model, the worse the predictive capability of just tweets. Therefore, we suggest that a combination disease-specific model -where the case counts of a disease are updated periodically along with the continuous monitoring of lexicon-based tweets plus or minus the time from disaster-would produce the best results.

Overall, the use of disease/sick lexicon-filtered tweets as a predictor of disease in the Philippines seems promising. Due to the consistency and rise of Twitter within the country, it would be informative to repeat analysis on more recent years to confirm the top method for prediction.

## Data Availability

Upon request, social media counts per region can be made available as well as disease count data.

## Data Availability Statement

Social media counts per region can be made available as well as disease count data.

